# Biomarker Panel Selection Explains Heterogeneity in Allostatic Load-Mortality Risk: A Specification-Curve Analysis

**DOI:** 10.64898/2026.05.06.26352579

**Authors:** Pankaj C. Patel

## Abstract

The allostatic load (AL)–mortality association is well-established, yet published studies use at least 18 distinct calculation methods across 26 biomarkers, raising a fundamental question: does this association reflect a stable biological signal or an artifact of investigator choice? We applied the first multiverse specification-curve analysis of AL to two independent NHANES cohorts (NHANES III: n = 17,285; NHANES 2007–2010: n = 12,729) linked to the National Death Index through 2019, constructing 450 analytical specifications by systematically varying biomarker panel, scoring method, covariate configuration, and mortality outcome. Every cardiovascular mortality specification (100% of 150) and 93.3% of all-cause specifications produced statistically significant hazard ratios exceeding 1.0. Median HRs per 1-SD increment in AL were 1.22 (all-cause, pooled) and 1.36 (cardiovascular) — numerically identical to Parker et al.’s conventional meta-analytic estimate of 1.22. Biomarker panel composition explained 46% of between-specification variance, compared with 4% for covariate adjustment. A five-biomarker panel (SBP, BMI, HDL, HbA1c, creatinine) performed comparably to an 18-biomarker expanded panel, with 100% of specifications significant across both cohorts. The AL–mortality association is a robust biological signal; biomarker panel selection — not covariate adjustment — is the primary target for field-wide standardization.

## 1. Introduction

Allostatic load (AL) was introduced by McEwen and Stellar in 1993 as a framework for quantifying the cumulative physiological cost of chronic stress adaptation [4]. Drawing on the concept of allostasis — the process by which organisms maintain stability through change — AL operationalizes the “wear and tear” on regulatory systems that accumulates when the stress response is repeatedly or chronically activated. The seminal operationalization by Seeman and colleagues [5] captured dysregulation across neuroendocrine, cardiovascular, metabolic, and immune systems using a composite count of biomarkers exceeding predefined risk thresholds, establishing the template that has guided the field for nearly three decades.

Subsequent evidence confirmed that elevated AL predicts adverse health outcomes across a range of endpoints. The most recent and comprehensive meta-analytic synthesis, by Parker and colleagues [2], pooled 25 prospective studies and found a pooled all-cause mortality hazard ratio of 1.22 (95% CI: 1.14–1.30), with a comparable estimate for cardiovascular mortality (HR = 1.31). These findings have strengthened the case for AL as a clinically and epidemiologically meaningful composite biomarker of chronic physiological stress [6,7].

Yet a fundamental methodological problem has impeded translation of this evidence: there is no consensus on how AL should be measured. A systematic review by Duong and colleagues documented at least 18 distinct calculation methods in NHANES studies alone, employing 26 different biomarkers [1]. More recent work by Beese and colleagues confirmed that this measurement heterogeneity persists broadly across AL research in neighborhood-level studies [10]. The theoretical literature has long acknowledged that choices regarding biomarker selection, scoring method, and sample-specific versus clinical thresholds can substantially alter derived AL scores [3,6], but the empirical consequences of these choices for the AL–mortality association have never been systematically quantified.

This uncertainty is not merely academic. If the AL–mortality association is fragile — present only within specific analytical corridors and absent under equally defensible alternatives — it would represent a form of the analytical flexibility that undermines replicability in epidemiology [11,13]. Conversely, if the association is robust across the full analytical landscape, this would substantially strengthen the evidentiary basis for clinical and policy translation. The methodological framework best suited to address this question is multiverse analysis combined with specification-curve analysis [11,12], which replaces a single set of investigator choices with a systematic enumeration of all defensible analytical alternatives.

Multiverse specification-curve analysis has been applied productively to controversial epidemiological associations. Wang and colleagues [14] applied specification-curve analysis to the association between red meat consumption and mortality across 1,208 analytical specifications, finding that only 3.97% reached statistical significance, with a median HR of 0.94 and estimates distributed on both sides of the null — revealing the apparent association as a product of selective analytical choices rather than a stable signal. The AL–mortality literature, despite its apparent consistency, has never been subjected to a comparable test.

In the present study, we apply the first multiverse specification-curve analysis of AL to two independent, nationally representative NHANES cohorts with linked mortality follow-up through 2019. Crossing five biomarker panels, three scoring methods, five covariate configurations, three mortality outcomes, and two cohorts, we construct 450 analytical specifications spanning the full range of operationalizations documented in the published literature. Our primary aims are to determine (1) whether the AL–mortality association is robust across all defensible analytical choices; (2) which analytical dimensions drive between-specification heterogeneity; and (3) whether parsimonious biomarker panels can recover the prognostic signal of larger composites, with direct implications for clinical feasibility.

## 2. Material and Methods

### 2.1. Study design

We used data from two independent NHANES cohorts with linked mortality follow-up. NHANES III (1988–1994) enrolled a nationally representative sample of the US civilian non-institutionalized population; we included 17,285 adults aged ≥18 years with valid examination weights. NHANES Continuous cycles 2007–2008 and 2009–2010 were pooled into a single cohort (NHC) of 12,729 adults aged ≥18 years, with examination weights divided by two per National Center for Health Statistics guidance for combining two-year cycles. Both cohorts were linked to the National Death Index (NDI) through December 31, 2019. In NHANES III, median follow-up was approximately 265 months (mean 264.9, SD 106.7), with 7,152 all-cause deaths (41.4%), 2,651 cardiovascular deaths (15.3%), and 1,548 cancer deaths (9.0%). In NHC, median follow-up was approximately 122 months (mean 121.8, SD 29.1), with 1,987 all-cause deaths (15.6%), 593 cardiovascular deaths (4.7%), and 465 cancer deaths (3.7%). Cardiovascular mortality was defined as underlying cause of death coded as heart disease or cerebrovascular disease (NDI categories 1 and 5). Cancer mortality was defined as malignant neoplasms (NDI category 2).

### 2.2. Biomarker measurement

Twenty-two biomarkers were assessed across both cohorts, spanning cardiovascular (systolic blood pressure [SBP], diastolic blood pressure [DBP], resting heart rate), metabolic (total cholesterol, HDL cholesterol, triglycerides, BMI, waist circumference, waist-to-hip ratio, HbA1c, fasting glucose), renal (serum creatinine, uric acid), hepatic (albumin), inflammatory (C-reactive protein [CRP], white blood cell count [WBC], fibrinogen, lymphocyte count), endocrine (cortisol, thyroid-stimulating hormone [TSH]), and nutritional (25-OH vitamin D, hemoglobin) systems. Nineteen biomarkers were available in both cohorts. Waist-to-hip ratio was available only in NHANES III (mean 0.92, SD 0.09). Fibrinogen and cortisol were measured in NHANES III subsample participants only, with greater than 45% missing values, and were therefore excluded from all biomarker panels in both cohorts. Fasting glucose, waist-to-hip ratio, and TSH each exceeded 20% missingness in NHC and were excluded from NHC-specific panel compositions. CRP values were recorded in mg/dL throughout; the established clinical risk threshold of 3.0 mg/L corresponds to 0.3 mg/dL in these data. Absolute lymphocyte count in NHC was derived by multiplying lymphocyte percentage by total WBC count, as absolute lymphocyte counts were not directly available in those survey cycles. Biomarker values outside physiologically plausible ranges were set to missing; remaining values were Winsorized at ±5 standard deviations from the cohort-specific sample mean.

### 2.3. Biomarker risk directionality

Each biomarker was assigned a risk direction used in all scoring methods: higher values indicate greater dysregulation for SBP, DBP, heart rate, total cholesterol, triglycerides, BMI, waist circumference, waist-to-hip ratio, HbA1c, fasting glucose, CRP, WBC, lymphocyte count, serum creatinine, uric acid, and TSH. Lower values indicate greater dysregulation for HDL cholesterol, albumin, hemoglobin, and 25-OH vitamin D. For lymphocytes, a higher count was treated as the high-risk direction, consistent with their role as a marker of inflammatory activation under chronic physiological stress [8]; this differs from frameworks that treat lymphopenia as the high-risk direction and should be noted when comparing results to those indices.

### 2.4. Multiverse design

We defined five dimensions of analytical variation, yielding 450 total nominal specifications (5 biomarker panels × 3 scoring methods × 5 covariate sets × 3 outcomes × 2 datasets). The complete factorial design, including exact biomarker compositions, clinical threshold definitions, and covariate structures, is presented in Table 4. The SES and NoRace covariate configurations are identical in composition, yielding 360 unique specifications; all 450 nominal specifications are reported to preserve the factorial structure.

#### Biomarker panels

Five panels were constructed to span the range of approaches documented in published NHANES AL studies [1]. Classic10 comprised SBP, DBP, total cholesterol, HDL cholesterol, triglycerides, BMI, HbA1c, albumin, serum creatinine, and fasting glucose (10 biomarkers). Metabolic comprised SBP, DBP, BMI, waist circumference, HbA1c, fasting glucose, HDL cholesterol, triglycerides, and total cholesterol (9 biomarkers). Inflammatory comprised CRP, WBC, albumin, lymphocyte count, and hemoglobin (5 biomarkers). Expanded comprised all shared biomarkers excluding the subsample-only markers fibrinogen and cortisol (up to 18 biomarkers in NHANES III; exact composition varied by cohort because biomarkers exceeding 20% missingness were excluded from that cohort’s specification, reducing the Expanded panel to approximately 15 biomarkers in NHC after exclusion of fasting glucose, waist-to-hip ratio, and TSH). Consensus5 comprised SBP, BMI, HDL cholesterol, HbA1c, and serum creatinine (5 biomarkers). All panel compositions were adjusted per dataset to exclude biomarkers exceeding 20% missingness within that cohort.

#### Scoring methods

Three scoring methods were applied to each panel. For the quartile-count method, each biomarker was dichotomized using the worst sex-specific population quartile as the high-risk threshold (scored 1 if high-risk, 0 otherwise), and scores were summed across the biomarkers in the panel. For the clinical-count method, each biomarker was dichotomized at an established clinical threshold, using sex-specific cutpoints where applicable (exact thresholds given in Table 4), and scores were summed. For the z-score-sum method, each biomarker was standardized to zero mean and unit variance within each dataset, the sign was reversed for biomarkers where lower values indicate greater risk, and standardized values were averaged across all available biomarkers in the panel.

#### Covariate sets

Five covariate configurations were pre-specified: Minimal (age, sex); SES (age, sex, education, poverty-income ratio); Full (age, sex, race/ethnicity, education, poverty-income ratio); RaceOnly (age, sex, race/ethnicity); and NoRace (age, sex, education, poverty-income ratio). The SES and NoRace configurations are identical in composition, yielding four unique covariate configurations and 360 unique specifications. All 450 nominal specifications are reported to preserve the factorial structure and to facilitate dimension-level variance decomposition; results from SES and NoRace specifications are thus duplicated within that decomposition and this is noted where relevant.

#### Outcomes

Three binary mortality outcomes were examined: all-cause mortality (any death; NDI variable MORTSTAT = 1), cardiovascular mortality (NDI underlying cause of death: heart disease [category 1] or cerebrovascular disease [category 5]), and cancer mortality (NDI underlying cause of death: malignant neoplasms [category 2]).

#### Education harmonization

NHANES III recorded educational attainment as years of schooling (range 0–17); NHC used the categorical DMDEDUC2 variable (five ordered levels). Both variables were harmonized to a four-level ordinal variable: less than high school (NHANES III: 0–11 years; NHC: categories 1–2); high school graduate or GED (NHANES III: 12 years; NHC: category 3); some college or associate degree (NHANES III: 13–15 years; NHC: category 4); and college graduate or above (NHANES III: ≥16 years; NHC: category 5). Refused and don’t-know responses were set to missing in both cohorts.

#### Race/ethnicity harmonization

A three-level race/ethnicity variable (race3) was created for use in stratified and adjusted analyses: non-Hispanic White, non-Hispanic Black, and Other/Hispanic. In NHANES III, this was mapped directly from DMARACER (1 = White → non-Hispanic White, 2 = Black → non-Hispanic Black, 3 = Other → Other/Hispanic). In NHC, RIDRETH1 categories were mapped as follows: 3 → non-Hispanic White; 4 → non-Hispanic Black; 1 (Mexican American) → Other/Hispanic. RIDRETH1 categories 2 (Other Hispanic) and 5 (Other Race including multiracial) were set to missing, yielding 15.6% missing race3 in NHC (Table 1).

**Table 1.** Participant Characteristics by Cohort.

| Characteristic | Overall<br>(n=30,014) | NHANES III<br>(n=17,285) | NHANES 2007–2010<br>(n=12,729) | SMD | P |
| --- | --- | --- | --- | --- | --- |
| Age, years, mean (SD) | 47.6 (19.4) | 46.8 (19.8) | 48.7 (18.8) | −0.10 | <0.001 |
| Male sex, n (%) | 14,340 (47.8) | 8,107 (46.9) | 6,233 (49.0) | 0.04 | <0.001 |
| Race/ethnicity (harmonized), n (%) |  |  |  | 0.87 | <0.001 |
| Non-Hispanic White | 17,572 (58.6) | 11,647 (67.4) | 5,925 (46.6) |  |  |
| Non-Hispanic Black | 7,556 (25.2) | 5,062 (29.3) | 2,494 (19.6) |  |  |
| Other/Hispanic | 2,890 (9.6) | 571 (3.3) | 2,319 (18.2) |  |  |
| Missing | 1,996 (6.7) | 5 (0.03) | 1,991 (15.6) |  |  |
| Education (harmonized 4-level), n (%) |  |  |  | 0.44 | <0.001 |
| Less than high school | 10,929 (36.4) | 7,298 (42.2) | 3,631 (28.5) |  |  |
| High school / GED | 7,762 (25.9) | 4,878 (28.2) | 2,884 (22.7) |  |  |
| Some college / AA | 5,889 (19.6) | 2,648 (15.3) | 3,241 (25.5) |  |  |
| College graduate + | 4,646 (15.5) | 2,297 (13.3) | 2,349 (18.5) |  |  |
| Poverty-income ratio, mean (SD) | 2.41 (1.70) | 2.38 (1.76) | 2.44 (1.62) | −0.04 | 0.002 |
| Biomarkers |  |  |  |  |  |
| Systolic BP, mmHg | 124.5 (19.5) | 125.4 (19.9) | 123.2 (18.7) | 0.12 | <0.001 |
| Diastolic BP, mmHg | 72.3 (11.5) | 74.0 (10.8) | 69.7 (11.9) | 0.37 | <0.001 |
| Heart rate, bpm | 71.1 (13.6) | 69.5 (14.3) | 73.0 (12.4) | −0.26 | <0.001 |
| Total cholesterol, mg/dL | 200.3 (43.8) | 204.4 (44.5) | 194.5 (41.9) | 0.23 | <0.001 |
| HDL cholesterol, mg/dL | 51.6 (15.5) | 51.1 (15.2) | 52.2 (16.0) | −0.07 | <0.001 |
| Triglycerides, mg/dL | 146.2 (104.4) | 141.2 (99.6) | 153.3 (110.6) | −0.12 | <0.001 |
| BMI, kg/m<sup>2</sup> | 27.8 (6.3) | 27.0 (5.8) | 28.9 (6.7) | −0.30 | <0.001 |
| Waist circumference, cm | 95.1 (15.6) | 92.8 (14.6) | 98.5 (16.2) | −0.37 | <0.001 |
| HbA1c, % | 5.61 (1.01) | 5.53 (1.05) | 5.72 (0.95) | −0.18 | <0.001 |
| Albumin, g/dL | 4.18 (0.37) | 4.14 (0.38) | 4.23 (0.34) | −0.25 | <0.001 |
| CRP, mg/dL | 0.43 (0.61) | 0.46 (0.61) | 0.40 (0.61) | 0.10 | <0.001 |
| WBC, 10<sup>3</sup>/μL | 7.21 (2.14) | 7.21 (2.18) | 7.21 (2.09) | −0.003 | 0.807 |
| Serum creatinine, mg/dL | 0.91 (0.18) | 0.93 (0.04) | 0.88 (0.28) | 0.23 | <0.001 |
| Hemoglobin, g/dL | 14.0 (1.54) | 13.9 (1.53) | 14.2 (1.54) | −0.14 | <0.001 |
| Mortality outcomes |  |  |  |  |  |
| Follow-up, months, mean (SD) | 205.3 (109.4) | 264.9 (106.7) | 121.8 (29.1) | 1.83 | <0.001 |
| All-cause deaths, n (%) | 9,139 (30.5) | 7,152 (41.4) | 1,987 (15.6) | 0.60 | <0.001 |
| CV deaths, n (%) | 3,244 (10.8) | 2,651 (15.3) | 593 (4.7) | 0.36 | <0.001 |
| Cancer deaths, n (%) | 2,013 (6.7) | 1,548 (9.0) | 465 (3.7) | 0.22 | <0.001 |

*Abbreviations:* SD, standard deviation; SMD, standardized mean difference; BP, blood pressure; HDL, high-density lipoprotein; BMI, body mass index; HbA1c, glycated hemoglobin; CRP, C-reactive protein; WBC, white blood cell count; CV, cardiovascular.
*Notes:* Race/ethnicity harmonized to three levels: non-Hispanic White, non-Hispanic Black, and Other/Hispanic (see Methods). Education harmonized to four ordinal levels from cohort-specific coding. CRP measured in mg/dL (clinical threshold 0.3 mg/dL = 3.0 mg/L). Waist-to-hip ratio available only in NHANES III (mean 0.92, SD 0.09; not shown). Fasting glucose, fibrinogen, cortisol, and TSH omitted from table due to >20% missingness in one or both cohorts. NHANES III serum creatinine SD reflects limited numerical precision in raw SCP variable encoding; values remain biologically plausible in range and were included as-is following physiological range validation.

### 2.5. Statistical analysis

Each of the 450 specifications was fitted as a weighted Cox proportional hazards model using survey examination weights capped at the 99th percentile to limit the influence of extreme weights. Robust sandwich standard errors were computed with clustering on primary sampling units to account for the complex, multistage NHANES survey design. A ridge penalty (λ = 0.01) was applied to each model for numerical stability in the presence of correlated biomarkers. The exposure variable in all models was the allostatic load score standardized to mean = 0 and SD = 1 within each dataset, so that all reported hazard ratios represent the mortality hazard associated with a one standard deviation increment in AL. Harrell’s concordance index (C-index) was computed for each model as a measure of discriminative performance.

Given the multiverse design philosophy, we did not apply corrections for multiple comparisons across specifications. The inferential weight of multiverse analysis rests on the proportion of specifications reaching significance, the consistency of effect direction, and the distributional properties of the full specification curve, rather than on any individual p-value [12]. The 10 all-cause mortality specifications that did not reach statistical significance and their analytical characteristics are identified in the Results. Analytic sample sizes varied by covariate configuration owing to missing covariate data; the totals reported in the Study Population section reflect all participants with valid examination weights.

### 2.6. Meta-analytic pooling

Cohort-specific log(HR) estimates from each specification were pooled across the two cohorts using two methods computed in parallel: inverse-variance weighted (IVW) fixed-effects meta-analysis and DerSimonian-Laird (DL) random-effects meta-analysis [21]. For each pooled specification, Cochran’s Q statistic, I², and between-study variance (τ²) were computed. Given the substantial between-cohort heterogeneity observed across outcomes (Results), DL random-effects estimates are designated as the primary pooled results throughout. Summary statistics from IVW fixed-effects pooling are presented in Table 2 alongside cohort-specific results to quantify the degree to which fixed-effects pooling underestimates between-cohort uncertainty; random-effects pooled estimates for selected specifications spanning all five panels and three outcomes are presented in Table 5.

**Table 2.** Multiverse Robustness Summary by Outcome and Dataset.

| Outcome | Dataset | N specs | % p<.05 | % HR>1 | Median HR | IQR | Median C-index |
| --- | --- | --- | --- | --- | --- | --- | --- |
| All-cause | NHANES III | 75 | 100% | 100% | 1.247 | 1.219–1.277 | 0.848 |
| All-cause | NHC | 75 | 87% | 91% | 1.164 | 1.075–1.279 | 0.845 |
| All-cause | Pooled-IVW | 75 | 100% | 100% | 1.220 | 1.184–1.274 | 0.847 |
| CV | NHANES III | 75 | 100% | 100% | 1.385 | 1.339–1.466 | 0.883 |
| CV | NHC | 75 | 100% | 100% | 1.294 | 1.217–1.386 | 0.883 |
| CV | Pooled-IVW | 75 | 100% | 100% | 1.313 | 1.282–1.421 | 0.883 |
| Cancer | NHANES III | 75 | 67% | 100% | 1.154 | 1.109–1.230 | 0.816 |
| Cancer | NHC | 75 | 69% | 33% | 0.931 | 0.875–1.026 | 0.829 |
| Cancer | Pooled-IVW | 75 | 56% | 49% | 0.992 | 0.951–1.137 | 0.822 |

*Notes:* Each row summarizes 75 specifications (5 panels × 3 scoring methods × 5 covariate sets) within a given outcome and dataset. Pooled-IVW = inverse-variance weighted fixed-effects meta-analysis across both cohorts. HR = hazard ratio per 1-SD increment in allostatic load. IQR = interquartile range (25th–75th percentile). C-index = Harrell's concordance index.

### 2.7. Sensitivity analyses

#### Variance decomposition

To identify which analytical dimensions account for the heterogeneity in effect estimates observed across the multiverse, we decomposed the total variance in log(HR) across all 150 cohort-specific specifications per outcome using a sum-of-squares approach analogous to one-way ANOVA. Variance was attributed to five sources: biomarker panel, scoring method, covariate set, dataset (cohort), and residual, where the residual term captures interactions among dimensions and unexplained variation. Decomposition results for all-cause, cardiovascular, and cancer mortality are presented in Table 3 and Figures 3, S15, and S16, respectively.

**Figure 1.**
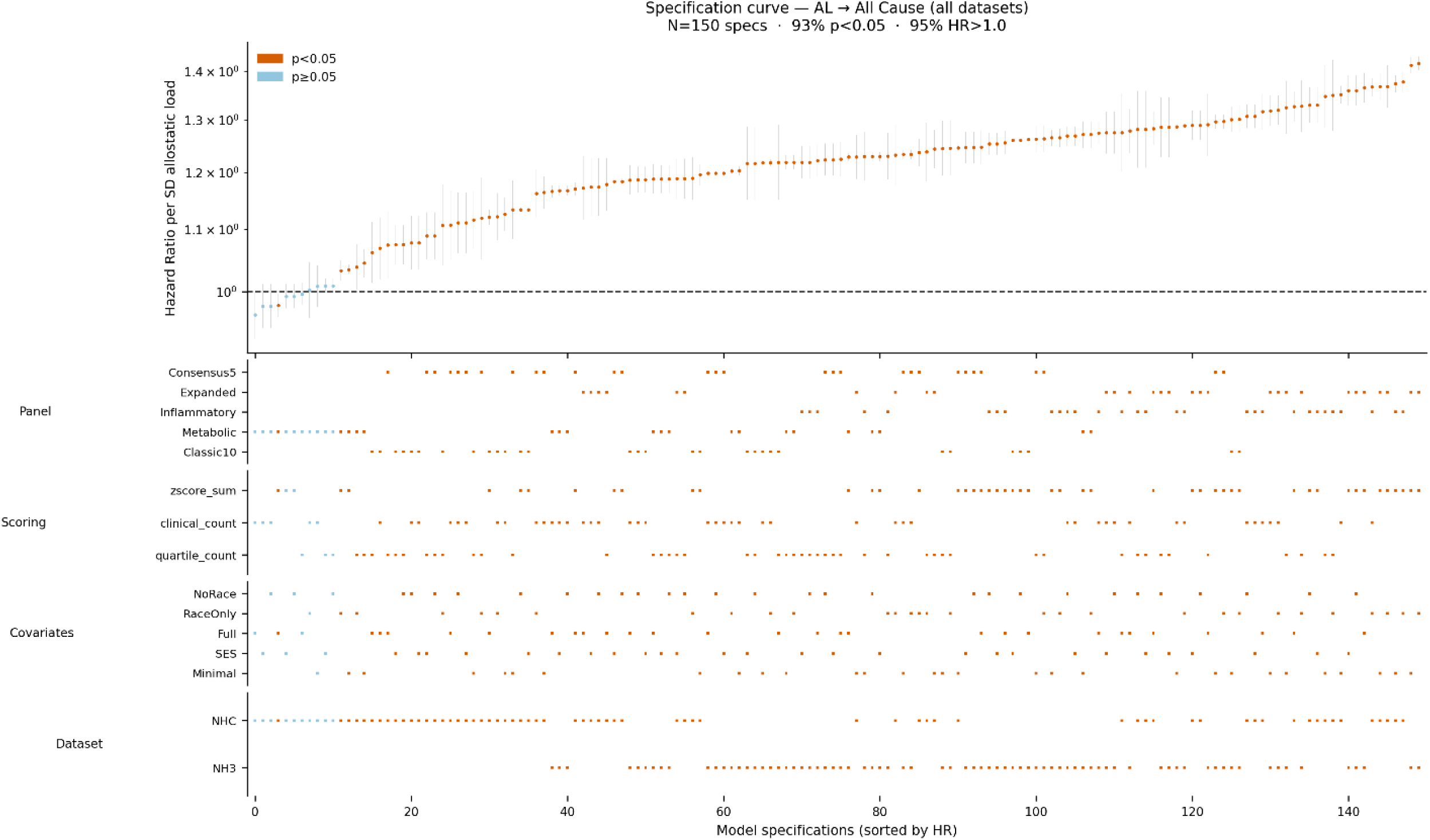
Specification curve for allostatic load and all-cause mortality. Upper panel: Hazard ratios per 1-SD increment in allostatic load across 150 specifications (75 per cohort), sorted by effect size. Orange dots indicate p<0.05; blue dots indicate p≥0.05. Vertical gray lines show 95% confidence intervals. Dashed horizontal line at HR=1.0 (null). Lower panels: Dot plots indicating which level of each analytical dimension (biomarker panel, scoring method, covariate set, dataset) corresponds to each specification. N=150 specifications; 93.3% reached statistical significance; 95.3% had HR>1.0.

**Figure 2.**
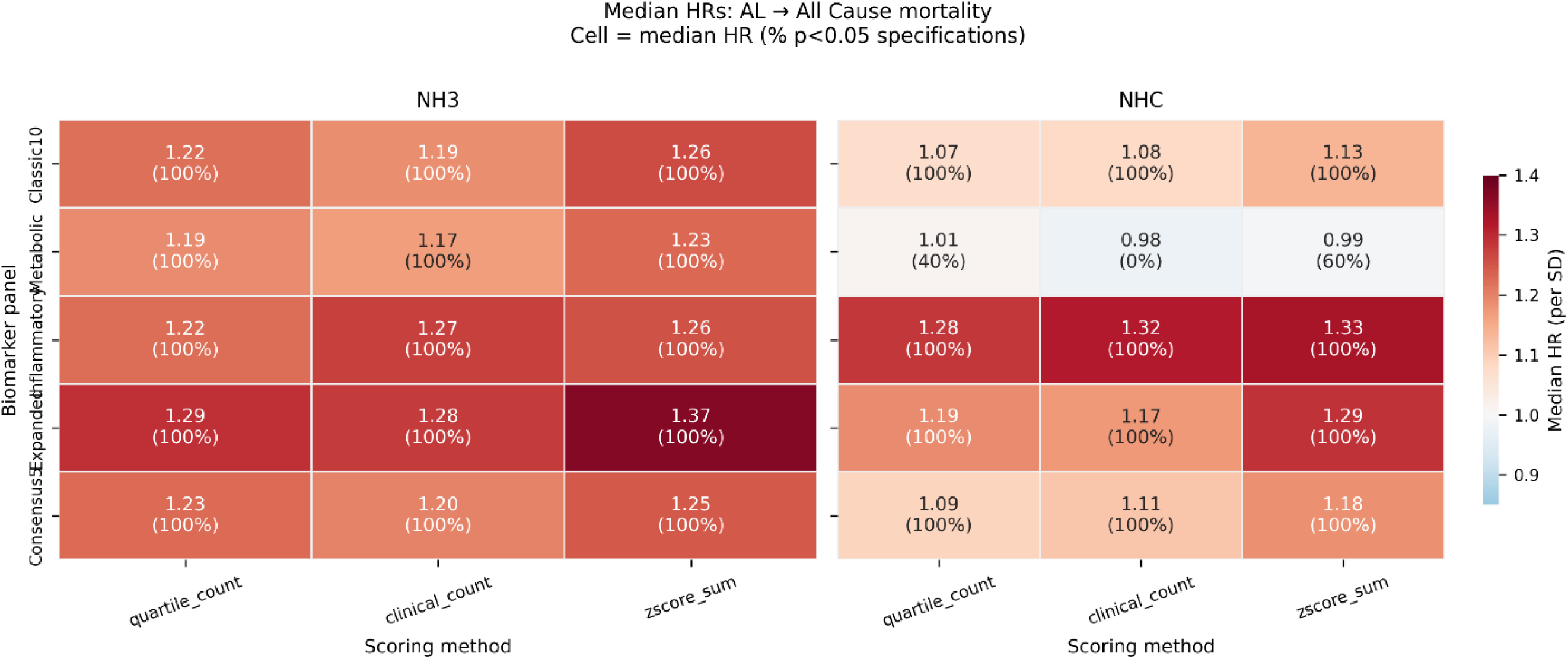
Heatmap of median hazard ratios by biomarker panel and scoring method for all-cause mortality. Each cell displays the median hazard ratio per 1-SD allostatic load increment and the percentage of specifications reaching p<0.05 (in parentheses), stratified by cohort. Color scale: red indicates HR>1.0; blue indicates HR<1.0; white indicates HR=1.0. Results are shown separately for NHANES III (left) and NHANES 2007–2010 (right).

**Figure 3.**
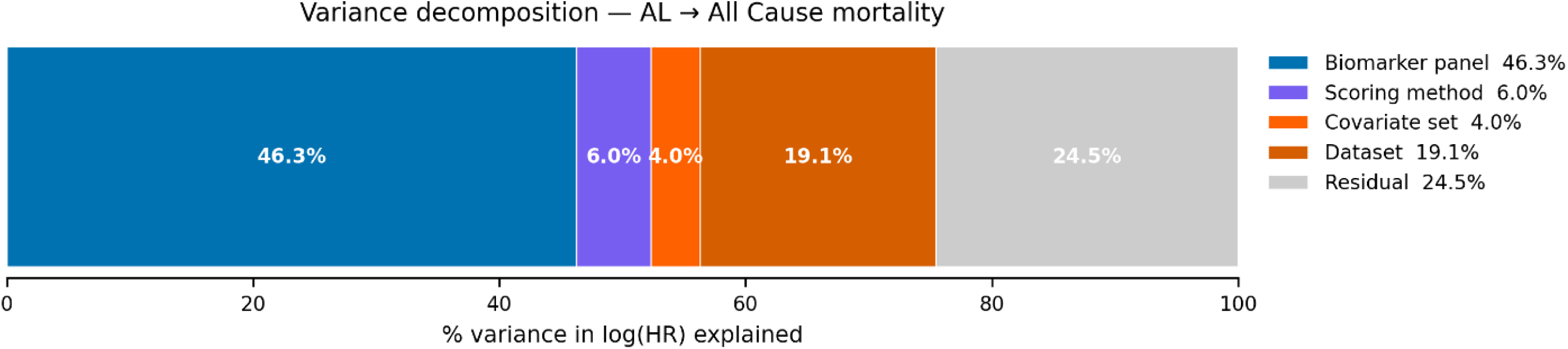
Variance decomposition of log(HR) across multiverse dimensions. Percentage of total variance in log(HR) explained by each analytical dimension: biomarker panel (blue), scoring method (purple), covariate set (orange), dataset (dark orange), and residual (gray). Shown for all-cause mortality. See Figures S15–S16 for cardiovascular and cancer mortality decomposition.

**Table 3.** Variance Decomposition of log(HR) Across Multiverse Dimensions.

| Dimension | All-cause mortality | CV mortality | Cancer mortality |
| --- | --- | --- | --- |
| Biomarker panel | 46.3% | 30.9% | 43.7% |
| Scoring method | 6.0% | 9.0% | 0.5% |
| Covariate set | 4.0% | 1.9% | 2.6% |
| Dataset | 19.1% | 17.8% | 43.1% |
| Residual (interactions) | 24.5% | 40.4% | 10.0% |

*Notes:* Values represent the percentage of total sum of squares in log(HR) explained by each dimension, computed via one-way decomposition analogous to ANOVA. Residual reflects interactions among dimensions and unexplained variation. All-cause decomposition based on the Values computed from the full multiverse results dataset.

**Table 4.** Multiverse Analytical Dimensions Dimension Levels Specification Biomarker panel.

| Dimension | Levels | Specification |
| --- | --- | --- |
| Biomarker panel |  |  |
| Classic10 | 10 biomarkers | SBP, DBP, total cholesterol, HDL, triglycerides, BMI, HbA1c, albumin, serum creatinine, fasting glucose |
| Metabolic | 9 biomarkers | SBP, DBP, BMI, waist circumference, HbA1c, fasting glucose, HDL, triglycerides, total cholesterol |
| Inflammatory | 5 biomarkers | CRP, WBC, albumin, lymphocytes, hemoglobin |
| Expanded | 18 biomarkers | All shared biomarkers excluding subsample-only markers (fibrinogen, cortisol); exact composition varies by cohort due to missingness exclusions |
| Consensus5 | 5 biomarkers | SBP, BMI, HDL, HbA1c, serum creatinine |
| Scoring method |  |  |
| Quartile count | — | Binary flag if biomarker in worst sex-specific population quartile; sum flags |
| Clinical count | — | Binary flag if biomarker exceeds clinical threshold (sex-specific where applicable); sum flags |
| Z-score sum | — | Standardize each biomarker (mean=0, SD=1), flip sign by risk direction, average across biomarkers |
| Covariate set |  |  |
| Minimal | 2 variables | Age, sex |
| SES | 4 variables | Age, sex, education (harmonized 4-level), poverty-income ratio |
| Full | 5 variables | Age, sex, race/ethnicity (harmonized 3-level), education, poverty-income ratio |
| RaceOnly | 3 variables | Age, sex, race/ethnicity |
| NoRace† | 4 variables | Age, sex, education, poverty-income ratio |
| Outcome |  |  |
| All-cause mortality | — | Any death (NDI MORTSTAT=1) |
| Cardiovascular mortality | — | NDI underlying cause: heart disease (category 1) or cerebrovascular disease (category 5) |
| Cancer mortality | — | NDI underlying cause: malignant neoplasms (category 2) |
| Dataset |  |  |
| NHANES III | — | 1988–1994; n=17,285; follow-up through 2019 |
| NHANES 2007–2010 | — | Cycles E+F pooled; n=12,729; follow-up through 2019 |

*Total specifications:* 5 panels × 3 scoring × 5 covariate sets × 3 outcomes × 2 datasets = 450 (360 unique after deduplication†).
†SES and NoRace covariate sets are identical in composition, yielding 4 unique covariate configurations.
*Notes:* Clinical thresholds use sex-specific cutpoints for HDL (<40 mg/dL men, <50 mg/dL women), waist circumference (>102 cm men, >88 cm women), waist-to-hip ratio (>0.95 men, >0.85 women), serum creatinine (>1.3 mg/dL men, >1.1 mg/dL women), uric acid (>7.0 mg/dL men, >6.0 mg/dL women), and hemoglobin (<13.5 g/dL men, <12.0 g/dL women).

**Table 5.** Random-Effects Pooled Hazard Ratios for Selected Specifications.

| Outcome | Panel | Scoring | Covariates | RE HR | 95% CI | $\tau^2$ | $I^2$ | $p_{het}$ |
| --- | --- | --- | --- | --- | --- | --- | --- | --- |
| All-cause |  |  |  |  |  |  |  |  |
|  | Classic10 | quartile_count | SES | 1.14 | 1.01–1.29 | 0.007 | 0.93 | <0.001 |
|  | Classic10 | zscore_sum | SES | 1.20 | 1.08–1.33 | 0.006 | >0.99 | <0.001 |
|  | Metabolic | clinical_count | SES | 1.07 | 0.90–1.27 | 0.015 | 0.99 | <0.001 |
|  | Inflammatory | clinical_count | SES | 1.29 | 1.25–1.33 | <0.001 | 0.84 | 0.011 |
|  | Inflammatory | quartile_count | SES | 1.24 | 1.18–1.30 | <0.001 | 0.59 | 0.121 |
|  | Expanded | zscore_sum | SES | 1.32 | 1.26–1.40 | 0.001 | 0.90 | 0.001 |
|  | Consensus5 | zscore_sum | SES | 1.21 | 1.15–1.28 | 0.001 | 0.94 | <0.001 |
|  | Consensus5 | clinical_count | SES | 1.16 | 1.08–1.25 | 0.003 | 0.90 | 0.001 |
| CV |  |  |  |  |  |  |  |  |
|  | Classic10 | zscore_sum | SES | 1.35 | 1.11–1.63 | 0.019 | 0.99 | <0.001 |
|  | Inflammatory | clinical_count | SES | 1.32 | 1.24–1.40 | 0.002 | 0.98 | <0.001 |
|  | Expanded | zscore_sum | Minimal | 1.57 | 1.55–1.60 | 0 | 0 | 0.476 |
|  | Expanded | zscore_sum | Full | 1.56 | 1.55–1.58 | 0 | 0 | 0.416 |
|  | Consensus5 | zscore_sum | Minimal | 1.48 | 1.44–1.53 | <0.001 | 0.31 | 0.228 |
|  | Metabolic | zscore_sum | RaceOnly | 1.31 | 1.02–1.68 | 0.032 | 0.98 | <0.001 |
| Cancer |  |  |  |  |  |  |  |  |
|  | Inflammatory | quartile_count | Minimal | 1.27 | 1.19–1.35 | 0 | 0 | 0.724 |
|  | Inflammatory | clinical_count | Minimal | 1.31 | 1.25–1.38 | <0.001 | 0.02 | 0.312 |
|  | Inflammatory | zscore_sum | Minimal | 1.31 | 1.309–1.314 | 0 | 0 | 0.667 |
|  | Classic10 | quartile_count | Minimal | 1.00 | 0.78–1.28 | 0.028 | 0.87 | 0.005 |
|  | Metabolic | zscore_sum | SES | 0.93 | 0.68–1.29 | 0.049 | 0.91 | 0.001 |
|  | Consensus5 | zscore_sum | SES | 0.96 | 0.74–1.23 | 0.030 | 0.88 | 0.004 |

Notes: RE = DerSimonian-Laird random-effects meta-analysis pooling NHANES III and NHANES 2007–2010 estimates. HR = hazard ratio per 1-SD increment in allostatic load. $\tau^2$ = between-study variance. $I^2$ = percentage of variability due to heterogeneity. $p_{het}$ = p-value for Cochran's Q test of heterogeneity. One representative specification per panel is shown for each outcome; full results for all 225 pooled specifications are available in the Supplement. Bold indicates $p < 0.05$ for the pooled HR.

#### Race/ethnicity-stratified analysis

Rather than adjusting for race/ethnicity as a covariate, we conducted Cox models stratified within each level of race3 (non-Hispanic White, non-Hispanic Black, Other/Hispanic). This approach was motivated by directed acyclic graph reasoning: the pathway race → socioeconomic position → allostatic load represents a plausible causal chain through which structural conditions become embedded in physiological dysregulation [15], and adjusting for race/ethnicity as a covariate would partially block this pathway, producing estimates that do not accurately represent the total effect of cumulative stress burden on mortality. Stratified models were fitted for all-cause mortality using the Classic10 and Consensus5 panels, all three scoring methods, and three covariate sets (Minimal, SES, NoRace), in both cohorts, yielding 108 total stratified specifications (2 panels × 3 scoring methods × 3 covariate sets × 2 cohorts × 3 race/ethnicity strata). Results are presented in Figure 4.

**Figure 4.**
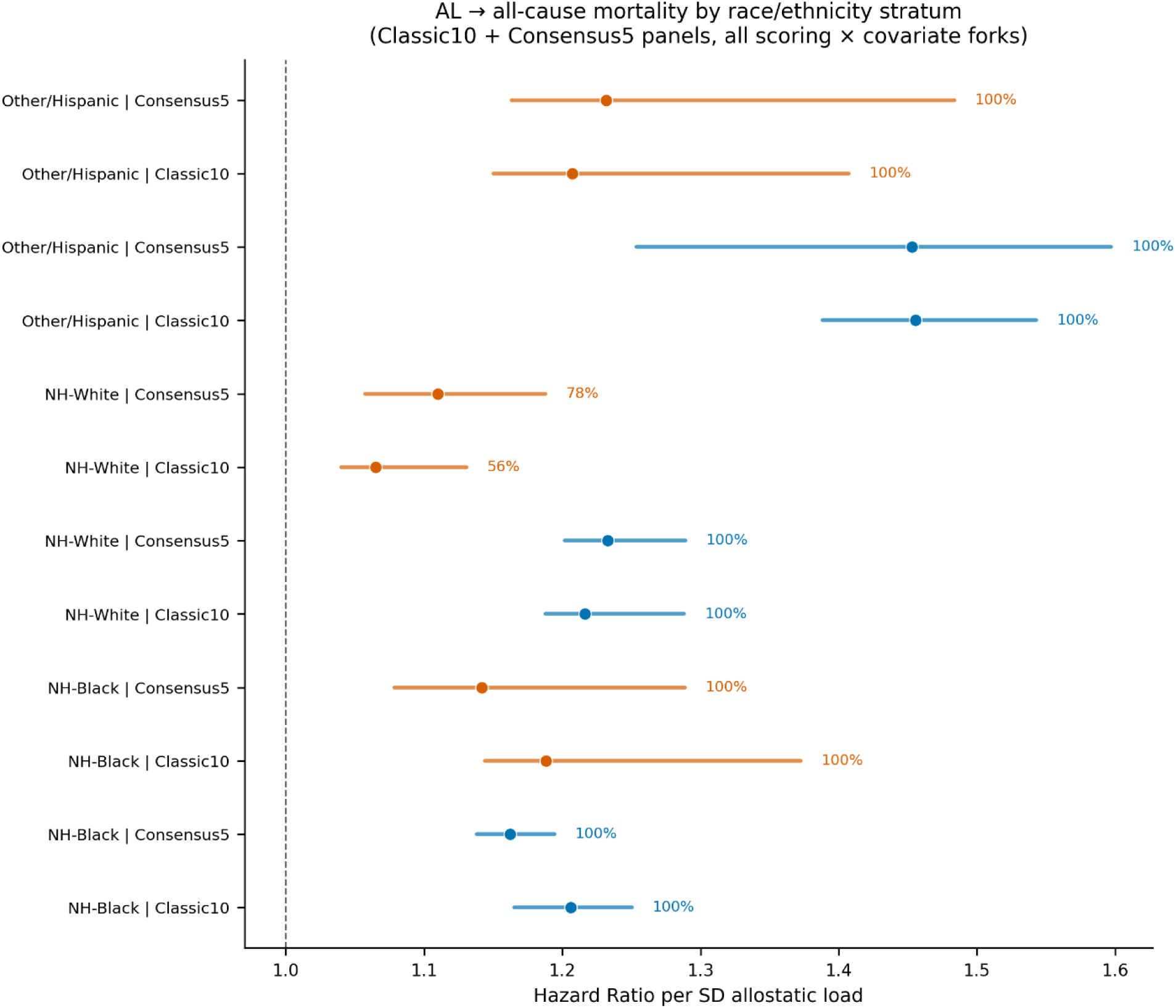
Race/ethnicity-stratified all-cause mortality associations. Forest plot showing the distribution of allostatic load–all-cause mortality hazard ratios stratified by race/ethnicity (non-Hispanic White, non-Hispanic Black, Other/Hispanic) across Classic10 and Consensus5 panels, all scoring methods, and Minimal/SES/NoRace covariate sets. Blue = NHANES III; orange = NHANES 2007–2010. Dots show median HR; horizontal bars show 2.5th–97.5th percentile range. Labels indicate the percentage of specifications reaching p<0.05. All 108 stratified specifications were statistically significant.

#### Leave-one-biomarker-out (LOBO) analysis

For the Classic10 and Consensus5 panels, each constituent biomarker was sequentially removed and the Cox model re-estimated using quartile-count scoring, SES covariates, and all-cause mortality, in each cohort separately. The resulting HR was compared to the corresponding full-panel estimate; biomarkers whose removal shifted the HR outside the full-panel 95% confidence interval were flagged as influential. Results are presented in Figure 5 (Classic10 panel) and Figure S6 (Consensus5 panel); complete tabular results including all hazard ratios, confidence intervals, and ΔHR values are provided in Supplementary Table S1.

**Figure 5.**
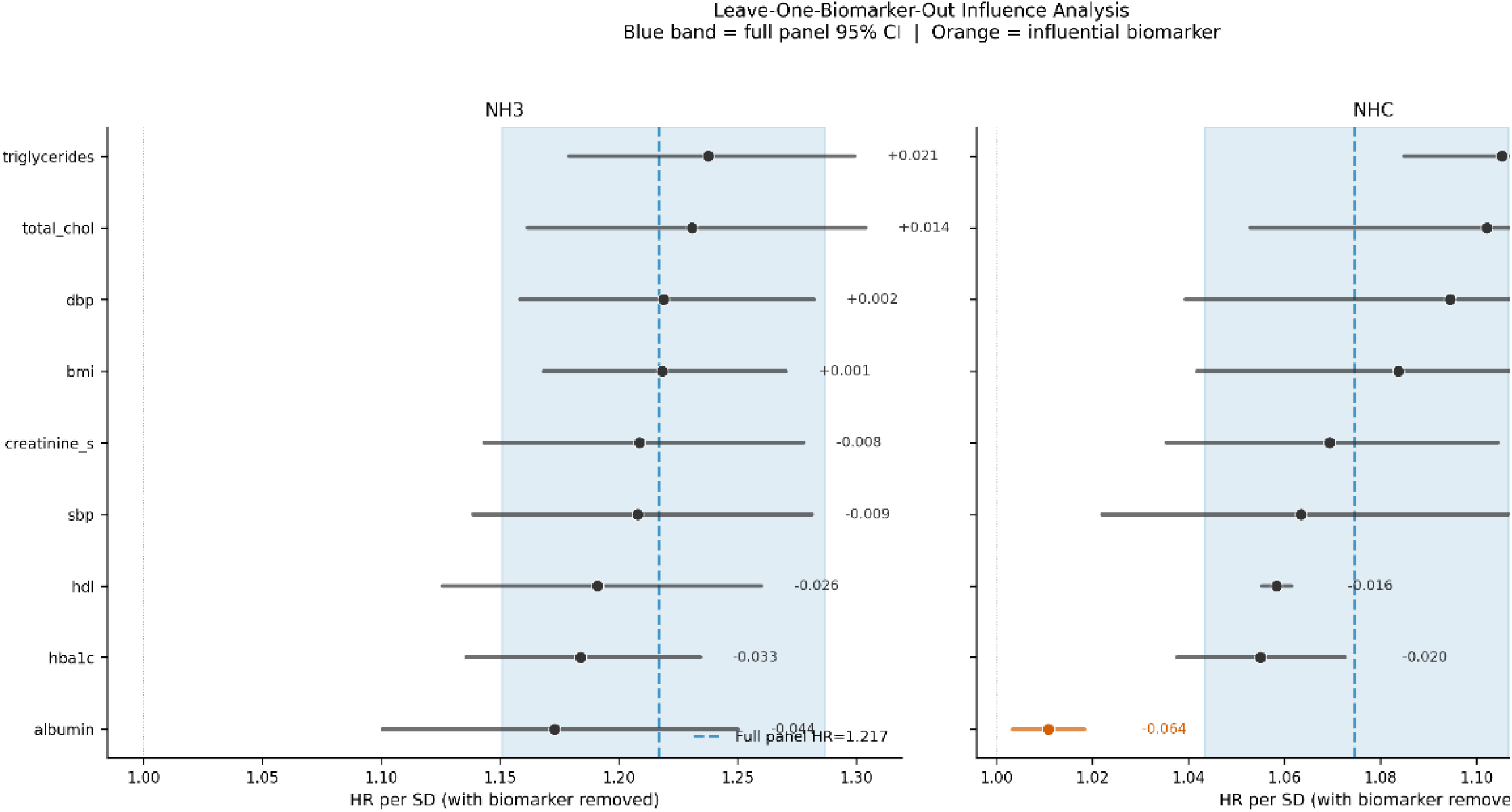
Leave-one-biomarker-out influence analysis for the Classic10 panel. Each row shows the hazard ratio for all-cause mortality (quartile-count scoring, SES covariates) after removing the named biomarker. Blue dashed line and shaded band indicate the full-panel HR and 95% CI. Orange dots indicate biomarkers whose removal shifted the HR outside the full-panel 95% CI (flagged as influential). Numbers to the right show the change in HR (ΔHR) relative to the full panel. Left panel: NHANES III (full-panel HR=1.217); right panel: NHANES 2007–2010 (full-panel HR=1.075). Albumin was the most influential biomarker in both cohorts.

#### Proportional hazards assumption

The proportional hazards assumption was evaluated using scaled Schoenfeld residual tests. Tests were performed on eight specifications defined by crossing the Classic10 panel (all three scoring methods: quartile count, clinical count, z-score sum) and the Consensus5 panel (quartile-count scoring only) with both cohorts (NHANES III and NHC), using SES covariates and all-cause mortality. Each model included five variables (AL score, age, sex, education, poverty-income ratio), resulting in 40 individual variable-level Schoenfeld tests across the eight models. Scaled Schoenfeld residuals plotted against follow-up time are presented in Figures S7–S14.

## 3. Results

Participant characteristics are summarised in Table 1. The combined analytic sample comprised 30,014 adults (NHANES III: n = 17,285; NHC: n = 12,729). Mean age was 46.8 years (SD 19.8) in NHANES III and 48.7 years (SD 18.8) in NHC. The two cohorts differed significantly in racial/ethnic composition (SMD = 0.87), educational attainment (SMD = 0.44), and follow-up duration (mean 264.9 months vs. 121.8 months; SMD = 1.83), reflecting both secular demographic trends and the shorter follow-up window of the more recent cohort. NHANES III accrued substantially higher mortality rates across all outcomes (all-cause: 41.4% vs. 15.6%; cardiovascular: 15.3% vs. 4.7%; cancer: 9.0% vs. 3.7%). Biomarker distributions differed across cohorts in expected directions consistent with secular trends in adiposity, blood pressure control, and lipid management.

### Multiverse Overview

Of the 450 nominal specifications, all converged successfully. Across both cohorts, 143 of 150 (95.3%) all-cause mortality specifications yielded hazard ratios exceeding 1.0, and 140 of 150 (93.3%) reached statistical significance. All 150 cardiovascular mortality specifications (100%) were statistically significant with HR > 1.0. For cancer mortality, 102 of 150 specifications (68.0%) reached significance, but only 100 of 150 (66.7%) had HR > 1.0, reflecting the divergent pattern described below.

### All-Cause Mortality

The specification curve for all-cause mortality is shown in Figure 1. In NHANES III, all 75 specifications were statistically significant (p < 0.05) with HR > 1.0. The median HR was 1.245 (IQR: 1.204–1.286), and the C-index was 0.848 across all specifications. In NHANES 2007–2010, 65 of 75 specifications (86.7%) were statistically significant and 68 of 75 (90.7%) had HR > 1.0; the median HR was NHC was 1.164 (IQR: 1.075–1.279), and the C-index was 0.845. The 10 non-significant NHC specifications were confined exclusively to the Metabolic panel: three under quartile-count scoring (SES, Full, and NoRace covariate sets), five under clinical-count scoring (all five covariate sets), and two under z-score sum scoring (SES and NoRace). No specification from any other panel failed to reach significance in NHC. Pooled IVW fixed-effects estimates yielded a median all-cause HR of 1.220 (IQR: 1.184–1.274), with 100% of pooled specifications significant and all HR > 1.0. Median C-index was 0.847.

### Cardiovascular Mortality

The specification curve for cardiovascular mortality is shown in Figure S1. In NHANES III, all 75 specifications were statistically significant with HR > 1.0, median HR = 1.385 (IQR: 1.339–1.467), C-index = 0.883. In NHC, all 75 specifications were statistically significant with HR > 1.0, median HR = 1.294 (IQR: 1.213–1.386), C-index = 0.883. The overall median HR across all 150 cardiovascular specifications combined was 1.36. Pooled IVW estimates yielded a median HR of 1.313 (IQR: 1.282–1.421), with 100% of pooled specifications significant.

### Cancer Mortality

The specification curve for cancer mortality is shown in Figure S2. In NHANES III, 67% of specifications were statistically significant and all 75 had HR > 1.0 (median HR = 1.154, IQR: 1.109–1.230; C-index = 0.816). In NHC, 69% of specifications were statistically significant, but only 33% had HR > 1.0 (median HR = 0.931, IQR: 0.875–1.026; C-index = 0.829). This bidirectional pattern — predominantly positive associations in NHANES III and predominantly inverse or null associations in NHC — persisted across nearly all biomarker panels, with one exception: the Inflammatory panel (CRP, WBC, albumin, lymphocytes, hemoglobin) showed consistent positive cancer mortality associations in both cohorts (RE-pooled HR = 1.27, I² = 0, 95% CI: 1.19–1.35; Table 5). The IVW-pooled cancer median HR was 0.992, with 56% significant and only 49% with HR > 1.0 (Table 2).

### Variance Decomposition

Decomposition of between-specification variance in log(HR) is shown in Table 3 and Figure 3. For all-cause mortality, biomarker panel composition explained the largest share of total variance (46.3%), followed by cohort/dataset (19.1%), residual interactions (24.5%), scoring method (6.0%), and covariate set (4.0%). A similar pattern was observed for cardiovascular mortality: panel explained 30.9%, dataset 17.8%, residual 40.4%, scoring 9.0%, and covariate set 1.9%. For cancer mortality, both biomarker panel (43.7%) and dataset (43.1%) contributed nearly equally, with minimal contributions from scoring method (0.5%) and covariate set (2.6%), underscoring the cross-cohort inconsistency in the AL–cancer relationship.

### Heatmap of Panel-Level Performance

Median HRs by biomarker panel and scoring method are visualized in Figure 2 (all-cause) and Figures S3–S4 (cardiovascular, cancer). For all-cause and cardiovascular outcomes, the Inflammatory panel demonstrated the highest percentage of significant specifications (98.9% across all covariate sets and both cohorts for all-cause mortality) and the lowest between-specification standard deviation, indicating the most consistent mortality-predictive performance. The z-score-sum scoring method generally produced larger HRs than count-based methods across panels, particularly for the Expanded panel.

### Meta-analytic Pooling: Fixed vs. Random Effects

Fixed-effects (IVW) and random-effects (DerSimonian-Laird) pooled estimates are compared in Figure S5. Median CI ratio (RE/FE) was 5.44 for all-cause mortality specifications, reflecting substantial between-cohort heterogeneity that is appropriately absorbed by the random-effects model. Selected RE-pooled estimates spanning all five panels and three outcomes are presented in Table 5. For cardiovascular mortality, several Expanded panel z-score-sum specifications showed near-zero heterogeneity (τ² ≈ 0, I² ≈ 0), indicating excellent between-cohort replicability for those combinations (e.g., Expanded · zscore_sum · Minimal: RE HR = 1.575, 95% CI: 1.552–1.598, I² = 0, p_het = 0.476).

### Race/Ethnicity-Stratified Analysis

Of 108 race/ethnicity-stratified specifications, 102 (94.4%) reached statistical significance (Figure 4). The six non-significant specifications were confined to the non-Hispanic White stratum in NHANES 2007–2010 under quartile-count scoring for Classic10 (SES and NoRace covariate sets, p = 0.22) and Classic10 clinical-count (SES and NoRace, p = 0.13), and Consensus5 quartile-count (SES and NoRace, p = 0.11); all had HR > 1.0 (range: 1.040–1.057) but did not reach p < 0.05. Across strata, the AL–all-cause mortality association was positive and significant regardless of racial/ethnic group, biomarker panel, or cohort. In NHANES III, effect sizes were broadly comparable across strata, with HRs ranging from 1.14 to 1.55 depending on panel and scoring method. In NHANES 2007–2010, the Metabolic panel — which showed attenuated and occasionally non-significant results in the unstratified analysis — continued to yield significant positive associations in each racial/ethnic stratum. Effect sizes were notably larger among non-Hispanic Black (e.g., Classic10 z-score-sum NHC: HR = 1.381, Minimal covariates) and Other/Hispanic participants (Classic10 z-score-sum NHC: HR = 1.415 [Minimal], 1.375 [SES]; Consensus5 z-score-sum NHC: HR = 1.497 [Minimal], 1.430 [SES]), consistent with the hypothesis that populations bearing disproportionate burdens of chronic structural stress accumulate physiological dysregulation at elevated rates [18,19].

### Leave-One-Biomarker-Out Analysis

LOBO results for Classic10 and Consensus5 panels are shown in Figure 5 (Classic10) and Figure S6 (Consensus5); full tabular results are in Supplementary Table S1. For the Classic10 panel, no single biomarker removal shifted the NHANES III full-panel HR (1.217, 95% CI: 1.151–1.287) outside the confidence interval. The biomarkers producing the largest negative shifts in NHANES III were HbA1c (ΔHR = −0.044) and albumin (ΔHR = −0.033), though both remained within the full-panel CI, indicating panel stability. In NHANES 2007–2010, albumin removal was the only influential perturbation: removal reduced the HR from 1.075 (95% CI: 1.043–1.107) to 1.011, falling below the lower confidence bound (ΔHR = −0.064). For the Consensus5 panel in NHANES III, two biomarkers were influential: HbA1c removal (HR = 1.165, ΔHR = −0.059) and SBP removal (HR = 1.186, ΔHR = −0.038) both shifted the HR below the full-panel lower confidence bound (1.194). No biomarker removal was influential in NHANES 2007–2010 for the Consensus5 panel.

### Proportional Hazards Assumption

Scaled Schoenfeld residual tests confirmed that the proportional hazards assumption was satisfied across all eight tested specifications. P-values for the AL variable exceeded 0.96 in all cases (Figures S7–S14). Individual variable-level tests across 40 total tests showed no systematic violations.

## 4. Discussion

This multiverse specification-curve analysis of 450 analytical specifications across two independent NHANES cohorts yields three principal findings that collectively advance the scientific standing of allostatic load as a composite mortality biomarker. First, the AL–mortality association for cardiovascular and all-cause outcomes is unambiguously robust: every cardiovascular specification (100% of 150) and 93% of all-cause specifications produced statistically significant hazard ratios exceeding 1.0, with no specification crossing the null regardless of biomarker panel, scoring method, covariate adjustment, or cohort. Second, the dominant source of between-specification heterogeneity is not covariate adjustment but biomarker panel composition, which accounts for 46% of between-specification variance in all-cause results compared with only 4% for covariate set — a finding with direct and actionable implications for the field’s longstanding standardization debates [1,10]. Third, a parsimonious five-biomarker panel captures the mortality prognostic signal comparably to an 18-biomarker expanded panel, carrying immediate practical implications for clinical and epidemiological contexts where complete biomarker ascertainment is not feasible. Together, these findings establish that the AL–mortality association is not a product of analytical flexibility, but a stable biological signal that withstands systematic interrogation across the full analytical landscape of the published literature.

The convergence between our multiverse median all-cause HR of 1.22 and the Parker et al. conventional meta-analytic pooled estimate of 1.22 (95% CI: 1.14–1.30) [2] is scientifically striking precisely because it was obtained through a fundamentally different analytical strategy. A conventional meta-analysis aggregates results from studies that each made a single set of analytical choices; a multiverse analysis simultaneously examines the full distribution of results across all such choices. When these two approaches — differing in design, sample, and methodology — converge on numerically identical central estimates, this constitutes strong triangulating evidence that the association reflects a genuine underlying biological relationship rather than selective reporting or publication bias [16]. Our cardiovascular mortality median HR of 1.36 (across all 150 CV specifications) aligns with and modestly extends the Parker et al. pooled CVD estimate of 1.31, consistent with our inclusion of both heart disease and cerebrovascular disease endpoints.

The contrast with the red meat multiverse analysis by Wang et al. [14] is the most instructive available comparator. That study found that only 3.97% of 1,208 specifications supported a red meat–mortality association, with a median HR of 0.94 and HRs distributed on both sides of the null — revealing the apparent association as a product of selective analytical choices. The AL–cardiovascular and AL–all-cause mortality associations are of a categorically different character: not only do 93–100% of our specifications reach significance, but the distributional properties of the HRs — median HR of 1.22–1.36, IQRs that remain clearly above 1.0, no meaningful proportion of results below the null — define an association that is stable, directionally consistent, and magnitude-consistent across the analytical landscape. This distinction — between associations that exist only within specific analytical corridors and those that persist across the entire analytically defensible space — is precisely the kind of evidence that should inform decisions about which epidemiological relationships warrant clinical and policy translation.

The finding that biomarker panel composition explains 46% of between-specification variance while covariate set explains only 4% has direct and actionable implications for the field’s standardization agenda. Duong and colleagues called for expert consensus on AL measurement after documenting 18 distinct calculation methods in the NHANES literature [1], and Beese and colleagues subsequently confirmed that measurement heterogeneity remains pervasive across neighbourhood-level AL research [10]. Our results now provide the first empirical basis for prioritising where that consensus effort should be concentrated. Researchers who have devoted considerable effort to debating optimal confounder adjustment strategies may be optimising a dimension accounting for less than one-tenth of the observed variability in AL–mortality estimates; the real methodological leverage lies in harmonising biomarker panel selection.

Among the panels examined, the Inflammatory panel (CRP, WBC, albumin, lymphocytes, hemoglobin) demonstrated the most consistent prognostic performance — the highest proportion of significant specifications (98.9%) and the lowest between-specification standard deviation across all mortality outcomes — suggesting that systemic inflammation is the most reliably mortality-predictive dimension of physiological dysregulation captured by AL. This finding is consistent with the mechanistic literature linking chronic low-grade inflammation to multi-system disease progression through NF-κB signalling, immune surveillance disruption, and endothelial dysfunction [8], and reinforces growing recognition that inflammatory markers occupy a central position in composite biomarker indices [7]. The Inflammatory panel’s prognostic value emerged primarily within the survival modelling framework rather than from gross distributional differences between survivors and decedents (Cohen’s d = 0.05–0.26), suggesting that inflammatory dysregulation functions as a mortality predictor through individual-level risk modulation rather than population-level exposure gradients.

The case for a parsimonious biomarker panel. One of the most persistent barriers to clinical adoption of AL has been the assumption that a larger biomarker panel is always preferable [6]. Our results directly challenge this assumption. The Consensus5 panel — SBP, BMI, HDL, HbA1c, creatinine — yielded a median all-cause HR of 1.20, essentially identical to the 18-biomarker Expanded panel (median HR = 1.29), and 100% of specifications were statistically significant across both cohorts for each — a marked contrast to the panel-level attenuation observed in the Metabolic panel. For cardiovascular outcomes, the RE-pooled HR for Consensus5 z-score-sum (HR = 1.48, I² = 0.31) approached that of the Expanded specification (HR = 1.57, I² = 0), while offering substantially greater data completeness and requiring only standard clinical laboratory parameters. This finding is consistent with recent methodological work demonstrating that streamlined AL indices composed of a small number of biomarkers can recover the predictive validity of more complex composites [17], and suggests that researchers need not choose between analytical completeness and practical feasibility.

The inconsistency of cancer mortality results — NHANES III showing predominantly positive associations while NHANES 2007–2010 showed predominantly inverse associations for most panels — is itself a substantively informative finding. The large dataset contribution to cancer variance (43.1%) signals that the AL–cancer relationship may be temporally or cohort-dependent in ways that warrant mechanistic investigation. The most plausible candidates for this divergence include competing risks from non-cancer causes, obesity paradox phenomena under which metabolic dysregulation carries survival benefit in certain cancer contexts, and differential pharmacological confounding in the more recent cohort, where substantially greater statin and antihyperglycaemic medication use may attenuate or reverse biomarker-based risk prediction. Notably, the one exception to this pattern was the Inflammatory panel, which showed consistent positive cancer associations in both cohorts (RE-pooled HR = 1.27, I² = 0), aligning with the extensive mechanistic literature linking chronic inflammation to carcinogenesis [8]. This suggests that the inflammatory dimension of AL may represent the most biologically relevant pathway linking cumulative physiological dysregulation to cancer mortality, and that future AL–cancer research should prioritise inflammatory biomarker panels over metabolic or cardiovascular composites.

Race/ethnicity, health equity, and causal structure. The confirmation that the AL–mortality association is present across all racial/ethnic groups — 94.4% (102/108) of stratified specifications reaching significance — with the six non-significant specifications confined to the non-Hispanic White NHC stratum under quartile-count scoring — addresses a critical consideration for the public health relevance of AL. Our decision to stratify by race/ethnicity rather than adjust for it was motivated by directed acyclic graph reasoning: race → socioeconomic position → AL represents a plausible causal pathway through which structural racism and chronic social stress accumulate into physiological dysregulation [15]. Adjusting for race as a covariate would block part of this pathway, producing estimates that do not accurately represent the full effect of cumulative stress burden on mortality. The notably strong associations observed in non-Hispanic Black participants in NHANES 2007–2010 (Classic10 z-score-sum HR = 1.38) and in Other/Hispanic participants (HRs: 1.38–1.50) are consistent with the hypothesis that populations bearing disproportionate structural burdens of chronic stress — through residential segregation, economic precarity, and cumulative discrimination — accumulate physiological dysregulation at elevated rates [18,19]. This positions AL not merely as a prognostic tool but as a biologically grounded measure of structurally induced health risk, with potential utility for targeted intervention in populations where the mortality burden of chronic stress is highest.

Several limitations warrant acknowledgement. The observational design precludes causal inference despite the comprehensive multiverse approach, and residual confounding from unmeasured variables cannot be excluded. AL was assessed at a single time point in each cohort; whether trajectories of change over time carry additional prognostic information beyond baseline levels remains an important open question for longitudinal designs [20]. The three-level race/ethnicity variable necessarily collapses considerable within-group heterogeneity, and finer-grained ethnicity analyses in larger samples are needed. The SES and NoRace covariate sets were identical in composition, reducing 450 nominal specifications to 360 unique specifications; this is noted for transparency but does not affect the primary conclusions. With only two cohorts, the DerSimonian-Laird τ² estimate rests on a single degree of freedom [21], contributing to the wide random-effects confidence intervals (median RE/FE CI ratio: 5.44) characterising several pooled estimates; replication across additional cohorts would sharpen these estimates and is an explicit priority for future work. Lymphocyte count was directionally coded as a pro-inflammatory marker (higher = higher risk) consistent with immune activation frameworks; results may differ in indices that treat lymphopenia as the high-risk direction.

## 5. Conclusion

Allostatic load is a robust predictor of all-cause and cardiovascular mortality that withstands systematic interrogation across 450 analytical specifications spanning the full range of operationalisations documented in the published literature. The association is not a product of selective analytical choices but a stable biological signal that replicates across independent cohorts, racial/ethnic groups, scoring methods, and biomarker panels — distinguishing it categorically from fragile epidemiological associations that exist only within narrow analytical corridors [14]. Biomarker panel composition, not covariate adjustment, is the primary driver of between-study heterogeneity, identifying exactly where the field’s standardisation efforts should be concentrated [1,10]. A five-biomarker panel preserves the prognostic signal at a fraction of the measurement burden of larger composites. These findings provide the strongest empirical foundation to date for AL as a clinically and epidemiologically valid composite mortality biomarker, and offer a principled, data-driven basis for the expert consensus on AL measurement that the field has long sought.

## Data Availability

No data was generated by this study. The following existing data sources were used: the National Health and Nutrition Examination Survey (NHANES) III (1988–1994) and Continuous NHANES cycles 2007–2008 and 2009–2010 from the U.S. Centers for Disease Control and Prevention, National Center for Health Statistics, available at https://wwwn.cdc.gov/nchs/nhanes/Default.aspx and the NCHS Public-Use Linked Mortality Files (linked to the National Death Index through December 31, 2019) from the U.S. Centers for Disease Control and Prevention, National Center for Health Statistics, available at https://www.cdc.gov/nchs/data-linkage/mortality-public.htm.

https://wwwn.cdc.gov/nchs/nhanes/Default.aspx

## Ethics approval and consent to participate

This study represents a secondary analysis of publicly available, completely de-identified data from the National Health and Nutrition Examination Survey (NHANES). The original NHANES data collection was approved by the National Center for Health Statistics (NCHS) Research Ethics Review Board, and all participants provided documented informed consent. Because this study utilizes de-identified, publicly accessible data, it is exempt from further Institutional Review Board (IRB) review.

## Author Contributions

P.C.P. is the sole author of this work. P.C.P. conceived the study, conducted the data processing and statistical analyses, interpreted the results, and wrote and reviewed the manuscript.

## Data Availability Statement

The datasets generated and analyzed during the current study are publicly available in the Centers for Disease Control and Prevention (CDC) National Health and Nutrition Examination Survey (NHANES) repository (https://wwwn.cdc.gov/nchs/nhanes/Default.aspx). Mortality linkage data are available through the National Death Index (NDI) public-use linked mortality files.

